# Genetic variants associated with longitudinal cognitive performance in older breast cancer patients and controls

**DOI:** 10.1101/2022.09.12.22279861

**Authors:** Kelly N. H. Nudelman, Kwangsik Nho, Michael Zhang, Brenna C. McDonald, Wanting Zhai, Brent J. Small, Claire E. Wegel, Paul B. Jacobsen, Heather S. L. Jim, Sunita K. Patel, Deena M. A. Graham, Tim A. Ahles, James C. Root, Tatiana M. Foroud, Elizabeth C. Breen, Judith E. Carroll, Jeanne S. Mandelblatt, Andrew J. Saykin, the Thinking and Living with Cancer (TLC) Study

## Abstract

**Background:** There have been no published genome-wide studies of the genetics of cancer- and treatment-related cognitive decline (CRCD); the purpose of this study is to identify genetic variants associated with CRCD in older female breast cancer survivors.

**Methods:** Analyses included white non-Hispanic breast cancer women with non-metastatic breast cancer aged 60+ (N=325) and age-, racial/ethnic group, and education-matched controls (N=340) with pre-systemic treatment and one-year follow-up cognitive outcomes. CRCD was assessed using longitudinal domain scores on neurocognitive tests of Attention, Processing speed, and Executive function (APE), and Learning and Memory (LM). Linear regression models of one-year cognition included an interaction term for SNP or gene SNP enrichment*cancer case/control status, controlling for demographic variables and baseline cognition.

**Results:** Cancer patients carrying minor alleles for two SNPs, rs76859653 (chromosome 1) in the hemicentin 1 (*HMCN1)* gene (p=1.624×10^−8^), and rs78786199 (chromosome 2, p=1.925×10^−8^) in an intergenic region had lower one-year APE scores than non-carriers and controls. Gene-level analyses showed the POC5 centriolar protein gene was enriched for SNPs associated with differences in longitudinal LM performance between patients and controls.

**Conclusion:** The SNPs associated with cognition in survivors, but not controls, were members of the cyclic nucleotide phosphodiesterase family, which play important roles in cell signaling, cancer risk, and neurodegeneration. These findings provide preliminary evidence that novel genetic loci may drive susceptibility to CRCD.

## INTRODUCTION

Improved early detection and treatments for breast cancer have greatly increased the number of survivors^1,2^. However, cancer and treatment-related cognitive decline (CRCD) has become an increasing concern^3^. CRCD is of particular concern in older survivors who constituted 62% of the 15.5 million cancer survivors in the United States in 2016, as age is a risk factor for cognitive decline and dementia, suggesting that this portion of the cancer survivor population might be more vulnerable to CRCD^1,3,4^.

Several studies have linked single nucleotide polymorphisms (SNPs) in candidate genes to CRCD. For example, studies have shown that the *APOE* e4 allele, the major risk factor for Alzheimer’s disease (AD), is associated with worse neurocognitive outcomes in some cancer patients^3,5-7^. More recently, there has been some evidence that *APOE* e2 may protect against CRCD in cancer survivors^8^. However, there have not been any published studies using a genome-wide analysis approach to identify loci associated with CRCD, and no genetic studies have focused on older survivors. Genome-wide investigation of CRCD genetic etiology could inform counseling and treatment of patients, as well as research on drugs targeting prevention and treatment of CRCD.

The primary objective of this study was to identify genetic variants showing different associations with longitudinal changes in neuropsychological domain scores for Attention, Processing speed, and Executive function (APE) or Learning and Memory (LM) in older breast cancer cases and non-cancer controls. The secondary objective was to identify genes enriched for variants showing different associations with longitudinal changes in APE and LM domains in cases and controls. These analyses aimed to identify variants and genes interacting with breast cancer diagnosis to affect risk of CRCD. This is the largest study of CRCD genetics published to date, and highlights the utility of this approach towards advancing the state of scientific knowledge in this field.

## MATERIALS AND METHODS

This study was a secondary analysis of specimens and data from the Thinking and Living with Cancer (TLC) study. TLC recruited participants from 13 oncology practices at or affiliated with six national sites: Georgetown University, Memorial Sloan Kettering Cancer Center, Moffitt Cancer Center, City of Hope Comprehensive Cancer Center, Hackensack University Medical Center, and Indiana University School of Medicine. All Institutional Review Boards approved the protocol (NCT03451383).

### Study Population

Participants were female breast cancer patients (stage 0-3) diagnosed at age 60+ years, and non-cancer controls frequency-matched on age and study site. Participants are followed prospectively with baseline pre-treatment/enrollment and then annual visits. For this study, baseline and one-year follow-up data were utilized for participants enrolled from 2010 to 2019. The TLC study is ongoing, and has been extensively described in other publications^3,7^. Briefly, participants were excluded for a history of stroke, head injury, major Axis I psychiatric disorders, neurodegenerative disorders, ever previously receiving chemotherapy or hormonal therapy, having had active treatment for cancer within the last five years prior to enrollment, or having a Mini-Mental State Examination score <24 or Wide Range Achievement Test-Fourth Edition (WRAT4) Word Reading score less than third grade level^9,10^. Additional eligibility for this analysis included having a biospecimen for GWAS testing and one-year cognitive data. To avoid bias from genetic ancestry and given the small number of minority participants in the study, genetic analyses were limited to White, non-Hispanic participants.

Of the 807 participants with imputed genetic data, 142 (95 cases, 47 controls) were missing clinical, demographic, or cognitive data, and were not included in the final analyses. These excluded participants were similar in age (mean=68.13, standard deviation=6.6), education (mean=15.1, standard deviation=2.3) and WRAT4 score (mean=109.8, standard deviation=14.2) compared to participants included in the analysis. A total of 665 cases (N=325) and controls (N=340) with imputed GWAS data and cognitive performance domain data were included in the analyses (See Figure 1 CONSORT Diagram).

**Figure 1.**
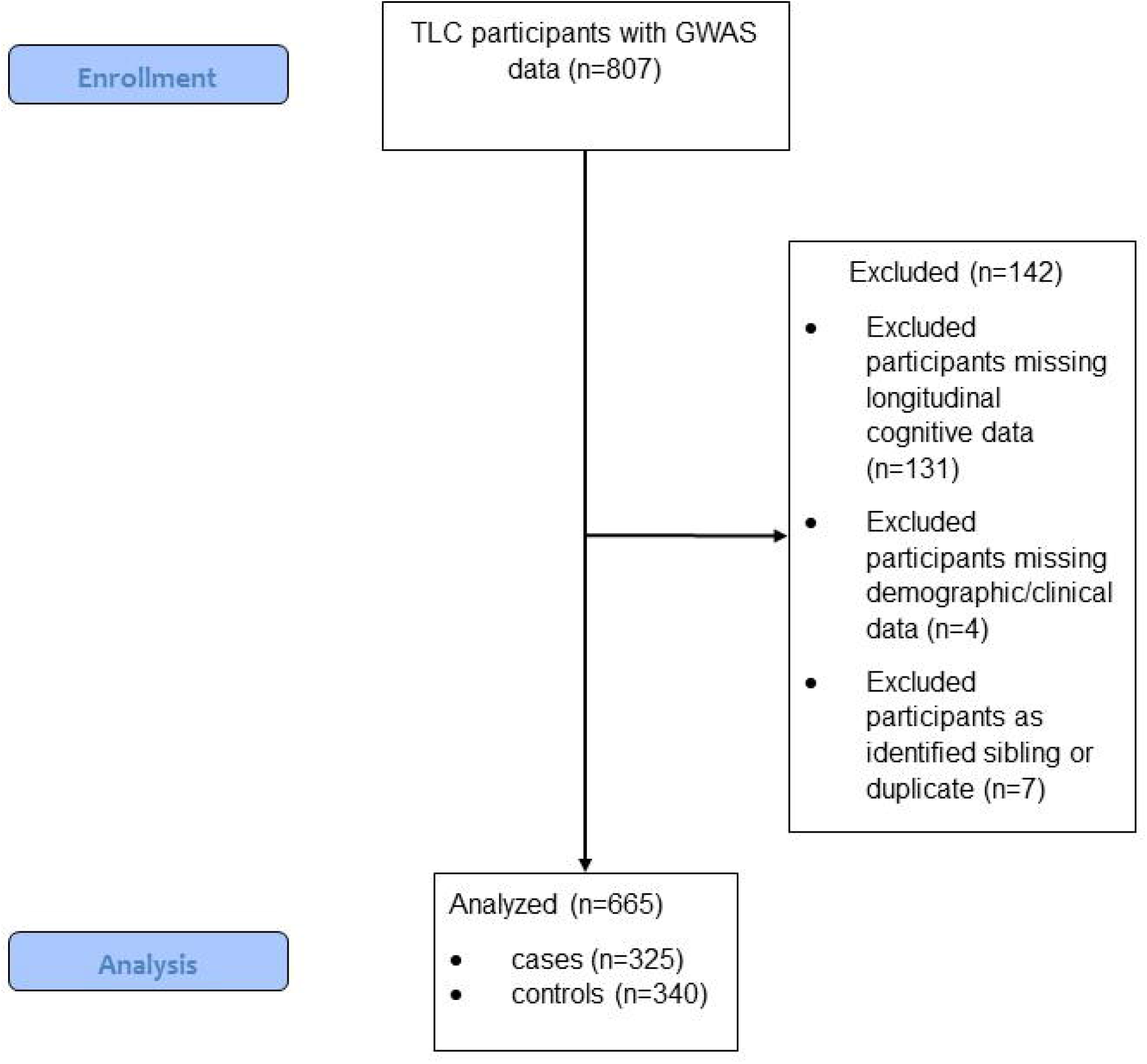
CONSORT Diagram. 807 White non-Hispanic participants had imputed GWAS data passing quality control. Of these, 142 were removed due to missing longitudinal data (131 for missing cognitive performance data, 4 for missing demographic/clinical information, and 7 for duplicate or first-degree sibling status). The final data set for analysis included 665 individuals, 325 cases and 340 controls.

### Data Collection

The baseline visit for TLC participants included collection of blood or saliva. In cases where a sample could not be collected at baseline, samples were collected at follow-up visits. Baseline assessments in patients were conducted following cancer-related surgery, but prior to initiation of chemotherapy, radiation, or hormone treatments.

Thirteen neuropsychological tests were administered at each visit to obtain data for two per protocol pre-specified cognitive domains: Attention, Processing speed, and Executive function (APE), and Learning and Memory (LM) (for more detail, see Supplementary Methods). Raw scores were standardized using the control group mean and standard deviation at baseline stratified by age and education. Standardized scores were then used to calculate z-scores for each domain for every participant, as described previously^11,12^. These domains were chosen because research has shown them to be relevant to CRCD^3^. WRAT4 reading scores were obtained at baseline^9^.

Collected demographic and clinical variables included age, years of education, collection site, race/ethnicity, and cancer and treatment information.

Saliva and/or blood samples were collected. Saliva samples were collected using Oragene kits (DNA Genotek, Kanata, Ontario, Canada); anticoagulated whole blood was collected with EDTA. Frozen EDTA samples and saliva samples at ambient temperature were shipped to Boston University or subsequently to the Indiana University Genetics Biobank to extract DNA, which was shipped frozen in three batches to the Children’s Hospital of Philadelphia, Center for Applied Genomics, where genome-wide association study (GWAS) assays were performed.

GWAS assays were performed using the Affymetrix Axiom Precision Medicine array (Thermo Fischer Scientific, Waltham, MA) for the first two batches and with the Illumina Global Screening Array v2 (Illumina, San Diego, CA) for the third batch. Microarray data were converted to PLINK format using Illumina GenomeStudio software (Illumina, Inc., San Diego, CA), and processed and quality-controlled with PLINK v1.9^13^. In total, 807 White non-Hispanic participants had genotype data passing quality control imputed with the haplotype Reference Consortium (HRC) panel using the Michigan Imputation Server^14,15^ (see Supplemental Methods for more details). The final data set included 7,661,137 SNPs, >10x the original number of SNPs obtained from genotyping. Of the 807 participants with imputed data, 131 participants were excluded for lack of one-year cognitive data, 4 were excluded due to missing covariates, and 7 were excluded from analysis as they were identified as duplicates or first degree siblings in the identity-by-descent analysis.

*Apolipoprotein E* (*APOE*) genotype was also obtained separately using TaqMan assays of rs429358 and r7412 on a Real-Time PCR System (Life Technologies, Carlsbad, CA), and/or Fluidigm genotyping with a custom-designed 96-SNP microarray (Fluidigm, San Francisco, CA).

### Statistical Methods and Analyses

Genetic data was analyzed in PLINK v1.9^13^. Primary analyses investigated the interaction of cancer case/control status with genotypes on one-year cognitive performance, controlling for baseline performance. Linear regression was used to predict one-year APE and LM scores based on the main effects of SNPs, group (cancer patient/non-cancer control), and SNP*group interaction, controlling for baseline cognitive scores, age, WRAT4 score, and recruitment site. Sensitivity analysis was performed to investigate the potential influence of *APOE* e4 carrier status; all models were run with/without *APOE* e4 as a covariate (data not shown).

SNP*case/control association analysis results were analyzed with The Functional Mapping and Annotation of Genome-Wide Association Studies (FUMA GWAS) program v1.3.6a^16^ (see Supplemental Methods for more information). For SNPs passing the genome-wide significance threshold (p<5×10^−8^), the most significant SNP from each locus showing an interaction with cancer group associated with cognitive performance was run in a general linear model in SPSS Statistics 25 (IBM SPSS Statistics 25, IBM Corp., Somers, NY), with interacting term cancer group, dependent factor APE one-year visit, and covariates baseline APE, age, and baseline WRAT4 to calculate marginal means for cancer case/control and carrier groups. Study site was entered as a fixed effect. The model was run with the interaction term SNP*cancer group as well as main effects for all terms, using a Type III sum of squares model including the intercept. Results included marginal means, standard deviations, and upper and lower bounds for the 95% confidence interval, as well as the F statistic and p-value for each SNP.

Secondary analyses of all 10,678 genes for enrichment of SNPs within a gene showing interaction with cancer case/control status associated with cognitive performance were also performed in FUMA using the MAGMA program v1.08^13,16-18^. Visualization of results in FUMA included generation of regional SNP plots using data from CADD^19^ and RegulomeDB^20^. For gene-level results, significance cut-off was p<5×10^−6^.

For lead SNPs at significant loci from the GWAS analyses, we also performed *post-hoc* testing of the SNPs for quantitative trait loci (QTLs) using the GTEx portal (gtexportal.org/) to investigate the functional consequences of each SNP on gene expression, splicing, and cell-specific regulation of gene expression^21^. For the intergenic SNP identified in the GWAS analysis, we investigated whether this locus was a predicted binding site for any transcription factors using JASPAR^22^, a database of transcription factor binding profiles.

## RESULTS

Participants were, on average 68 years old (range 61 to >90), and cases and controls had >15 years education (Table 1). Differences in education and WRAT4 scores between cases and controls were not clinically meaningful. There were no significant differences between women with breast cancer and controls for *APOE* e4 allele frequency.

**Table 1.** Demographics.

| Variable | Case<br>(N=325)* | Control<br>(N=340) | *p-value\*\** |
| --- | --- | --- | --- |
| Age, mean years (StDev) | 68.2 (5.7) | 67.9 (6.6) | *0.596* |
| Education, mean years (StDev) | 15.3 (2.1) | 15.7 (2.2) | **0.012** |
| WRAT4 score, mean (StDev) | 111.0 (15.8) | 113.7 (15.4) | **0.028** |
| Chemotherapy treatment<br>number (%) | 84 (25.8%) | - | - |
| Hormone therapy | 257 (79.1%) | - | - |
| *APOE* e4 carrier, number (%) | 81 (24.9%) | 85 (25.0%) | *1.000* |

StDev = standard deviation; WRAT4 = Wide Range Achievement Test-Fourth Edition Word Reading Test; *APOE* = apolipoprotein E
\*84 (25.8%) of the 325 cases were treated with chemotherapy between the baseline and one-year visits
\*\*p-values are italicized, with values <0.05 on ANOVA or Fischer's Exact two-sided test shown in bold

### GWAS and Gene Analyses

#### GWAS Analyses

Two loci, on chromosomes 1 (rs76859653, p=1.624×10^−8^) and 2 (rs78786199, p=1.925×10^−8^), were differentially associated with longitudinal APE performance in breast cancer cases compared to controls (see Figure 2A GWAS Manhattan plots for genome-wide analysis results, Figure 3 for plots of each SNP locus, supplementary Figure 1A for GWAS QQ plots). As shown in Figure 4 and Table 2, control individuals carrying minor alleles for either SNP have similar or greater APE one-year mean scores compared to non-carriers controlling for baseline scores. In contrast, cases carrying minor alleles for either of these SNPs have lower APE one-year mean scores than non-carriers controlling for baseline scores, suggesting that in cancer patients but not controls, carriers for either SNP have a greater risk for cognitive decline over time. The analysis of LM domain performance did not identify any SNPs of genome-wide significance (p<5×10^−8^, Figure 2B, supplementary Figure 1B).

**Figure 2.**
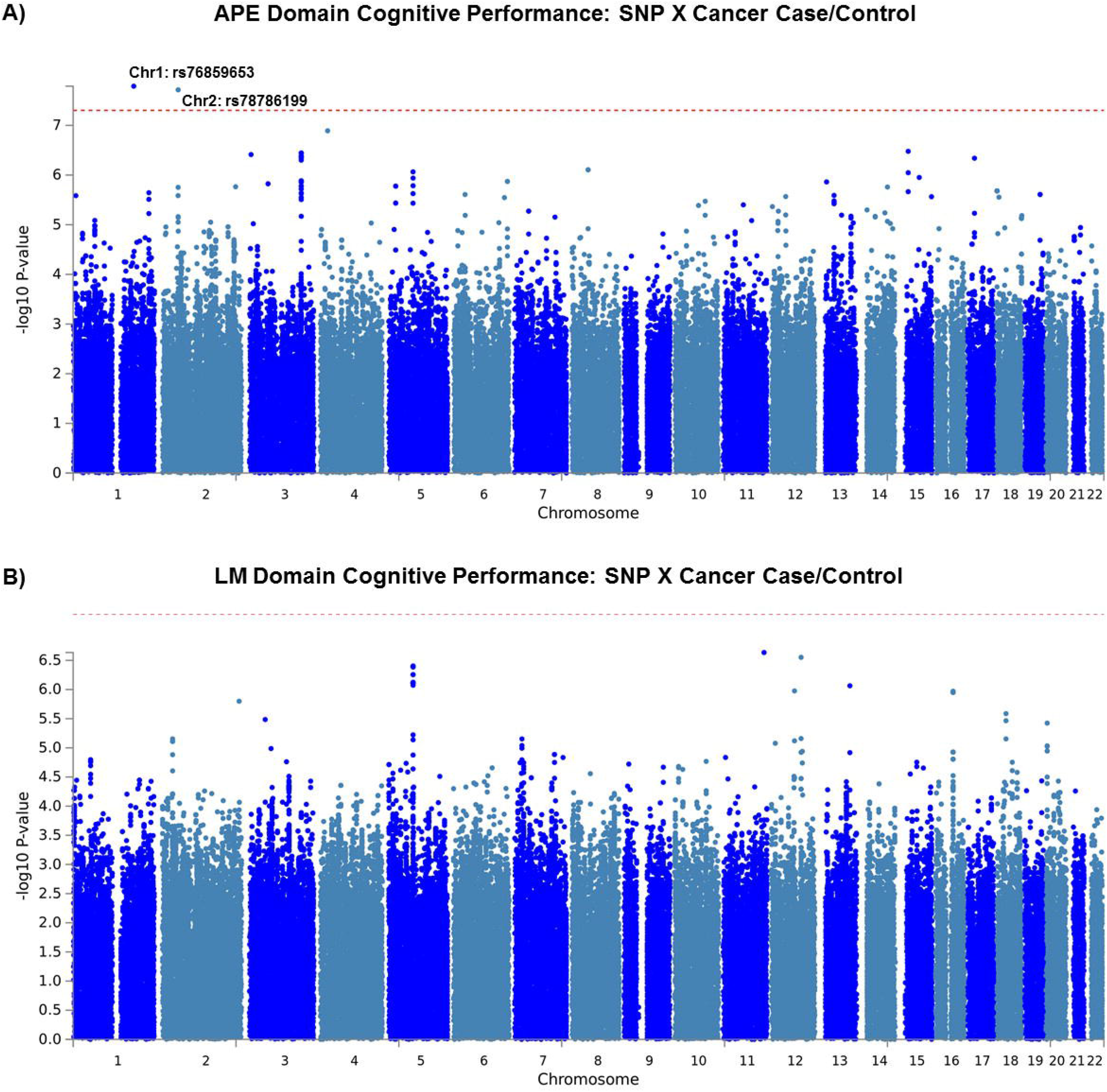
GWAS SNP*Cancer Interaction. A) Manhattan plot of GWAS SNP by group interaction associated with one-year follow-up (1Y) visit Attention, Processing speed, and Executive function (APE) cognitive domain score. GWAS genome-wide analysis of SNP*group (0/1) interaction with outcome of 1Y visit APE score, covarying for age, baseline WRAT4 score, site, and baseline APE score. Loci on chromosomes 1 (rs76859653, p=1.624×10^−8^) and 2 (rs78786199, p=1.925×10^−8^) have p-values of genome-wide significance (p<5×10^−8^). B) Manhattan plot of GWAS SNP by group interaction associated with 1Y visit Learning and Memory (LM) cognitive domain score. GWAS genome-wide analysis of SNP*group (0/1) interaction with outcome of 1Y visit LM score, covarying for age, baseline WRAT4 score, site, and baseline LM score. No loci attained genome-wide significance (p<5×10^−8^).

**Figure 3.**
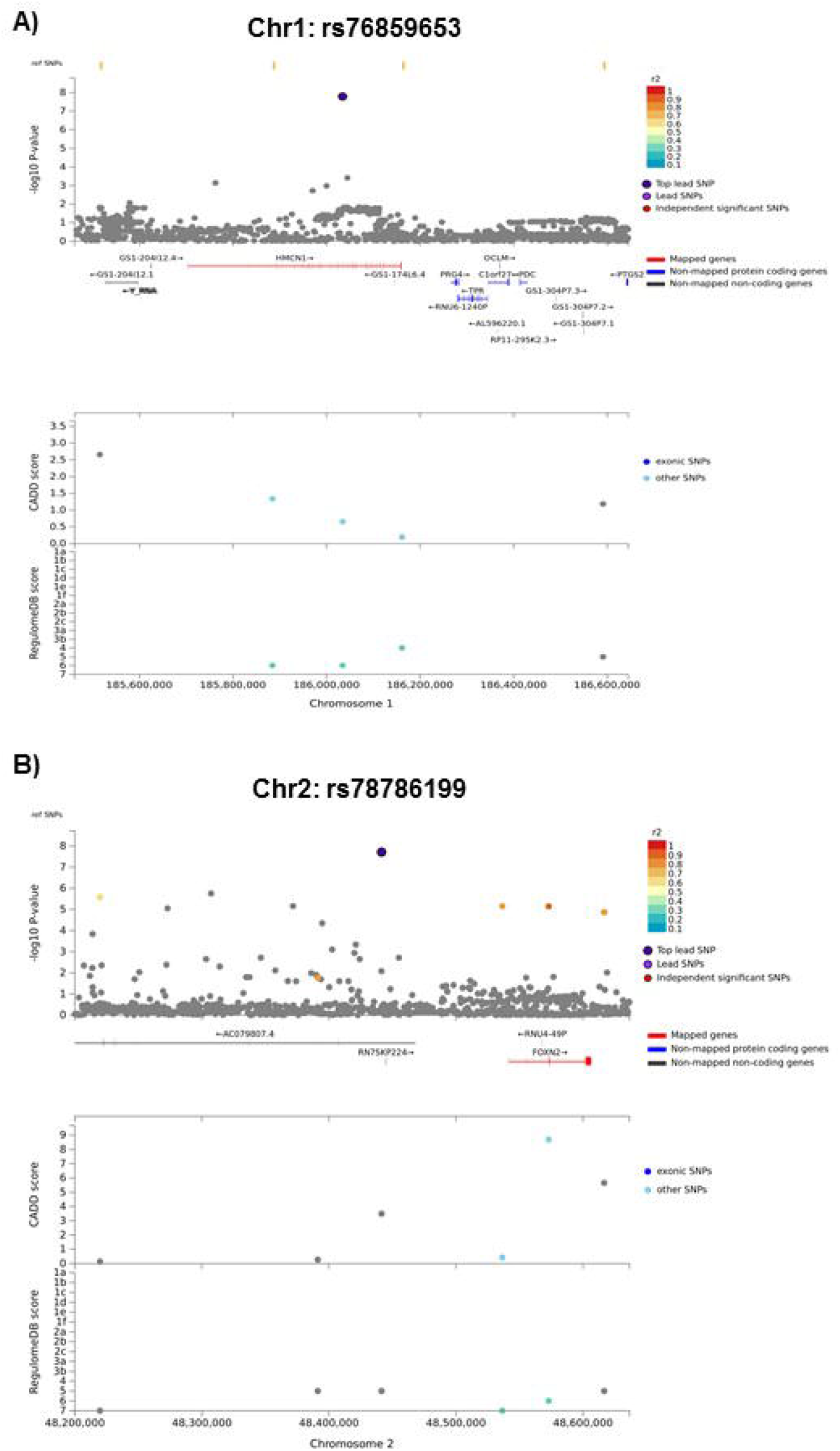
Regional Plots for Lead SNPs from APE GWAS Analysis. Top panel shows the SNPs adjacent to the lead SNP. Top SNP is colored navy; independent significant SNPs are color coded by r2 scale. SNPs that are not in linkage disequilibrium (LD) with the top lead SNP are shown in gray. ‘Ref SNPs’, displayed at the top of the plot, are SNPs that are in LD with the top SNP, but which do not have a p value because they were not included in the data. Mapped genes are shown in red. Y-axis shows the –log10 p-value of all graphed SNPs. The bottom panel shows exonic SNPs (dark blue) and other SNPs (light blue), graphed for CADD score (SNP deleteriousness) and RegulomeDB score (evidence for transcription factor function); increasing values for both indicate increasing likelihood of function. Location on the chromosome in base pairs is shown on the X-axis. A) Regional SNP Plot for rs76859653 on chromosome 1, a lead SNP significantly differently associated with APE cognitive domain performance at one-year follow-up (1Y) visit by group, covarying for age, baseline WRAT4 score, site, and baseline APE performance. B) Regional SNP Plot for rs78786199 on chromosome 2, a lead SNP significantly differentially associated with APE cognitive domain performance at 1Y visit by group, covarying for age, baseline WRAT4 score, site, and baseline APE performance.

**Figure 4.**
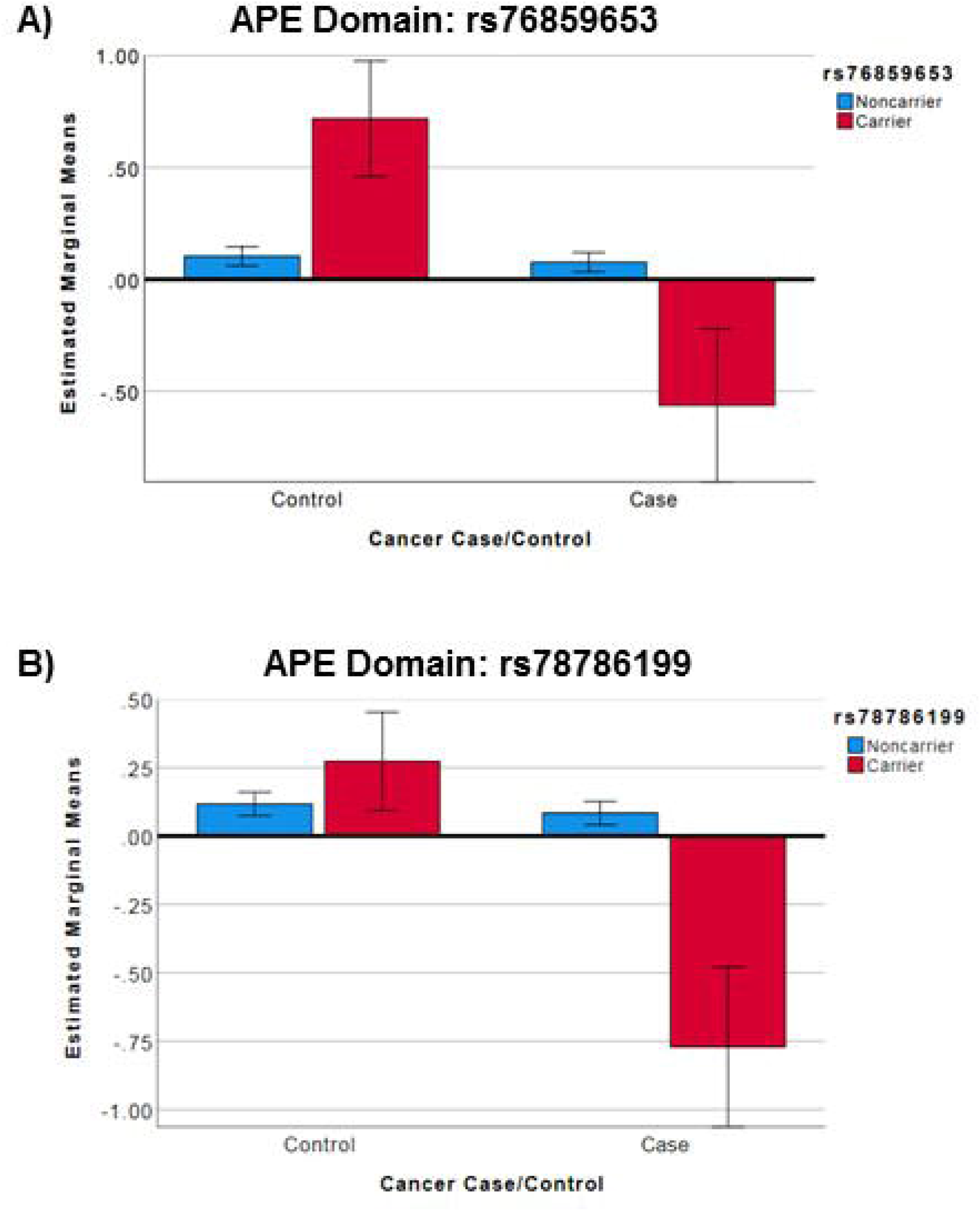
SNPs Differentially Associated with APE Score by Group. Boxplots depicting differences in APE score at one-year follow-up (1Y) visit (Y axis) by cancer case/control status (X-axis) and by SNP minor allele carrier status (red=carrier, blue=noncarrier). There were no individuals homozygous for either SNP; all carriers are heterozygous. Error bars indicate 95% confidence intervals. Covariates appearing in the model were evaluated at the following values: age = 68.05, WRAT4 = 112.33, baseline APE = 0.03, site 1 = 1.21, site 2 = 1.17, site 3 = 1.28, site 4 = 1.14, site 5 = 1.07. A) Results for rs7659653. There were 329 control noncarriers, 9 control carriers, 319 case noncarriers, and 5 case carriers. B) Results for rs78786199. There were 322 control noncarriers, 18 control carriers, 317 case noncarriers, and 7 case carriers.

**Table 2.** SNPs associated with Longitudinal Attention, Processing Speed, and Executive Function (APE) domain scores differentially in cancer cases vs. controls□.

| SNP | Group | SNP MA* | APE<br>Mean** (StE) | 95% CI<br>Lower<br>Bound | 95% CI<br>Upper<br>Bound | F | p<br>value** |
| --- | --- | --- | --- | --- | --- | --- | --- |
| rs76859653 | Control | 0 (N=329) | 0.10 (0.02) | 0.06 | 0.15 | 32.68 | <0.001 |
|  | Control | 1 (N=9) | 0.71 (0.13) | 0.46 | 0.97 |  |  |
|  | Case | 0 (N=319) | 0.08 (0.02) | 0.03 | 0.12 |  |  |
|  | Case | 1 (N=5) | -0.56 (0.17) | -0.91 | -0.22 |  |  |
| rs78786199 | Control | 0 (N=322) | 0.12 (0.02) | 0.07 | 0.16 | 32.27 | <0.001 |
|  | Control | 1 (N=18) | 0.27 (0.09) | 0.09 | 0.45 |  |  |
|  | Case | 0 (N=317) | 0.08 (0.02) | 0.04 | 0.13 |  |  |
|  | Case | 1 (N=7) | -0.77 (0.15) | -1.06 | -0.48 |  |  |

StE = Standard Error; CI = confidence interval.
\*0=homozygous for major allele, 1=heterozygous for minor allele; neither of the top SNPs have any homozygous minor allele genotypes in the data set.
\*\*Results from general linear models with individual significant SNPs identified from GWAS analysis (APE one-year score ~ SNP\*group + covariates); cognitive means for APE score calculated controlling for mean-centered covariates.

For rs76859653 and rs78786199, GTex analysis showed no significant QTLs for either SNP. While this analysis did not identify any genes with differential expression associated with these SNPs, investigation of intergenic rs78786199 with JASPAR showed that this SNP is within the region of predicted transcription factor binding sites for Zinc Finger Imprinted 3, Interferon Regulatory Factors 1, 4, 7, and 8, Signal Transducer and Activator of Transcription 2, and Zinc Finger Protein 317.

Analyses performed with *APOE* e4 carrier status as an additional covariate did not differ significantly (data not shown).

#### Gene Analyses

Gene analysis did not identify any genes enriched for variants significantly associated (p<5×10^−6^) with APE one-year score controlling for baseline when comparing cancer patients to controls (see Figure 5A for gene analysis results, supplementary Figure 2A for gene QQ plots). Of note, the most significant results for this analysis were phosphodiesterase 3A (*PDE3A*, p=5.77×10^−6^) and phosphodiesterase 4B (*PDE4B*, p=1.49×10^−5^), both of which are members of the cyclic AMP-specific cyclic nucleotide phosphodiesterase (PDE) family.

**Figure 5.**
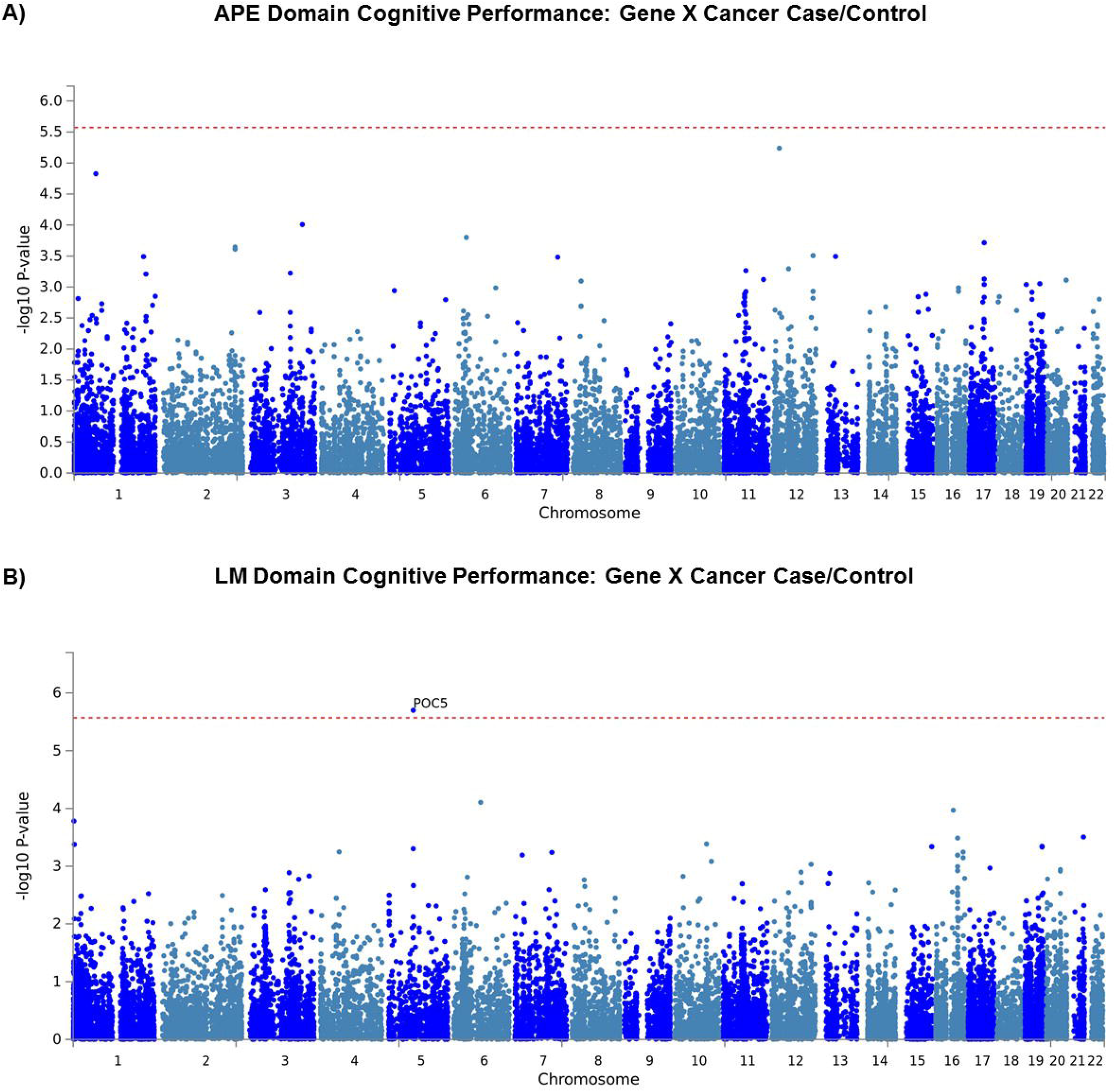
GWAS Gene*Group Interaction. A) Manhattan plot of TLC gene by group interaction associated with one-year follow-up (1Y) visit Attention, Processing speed, and Executive function (APE) cognitive domain score. GWAS genome-wide analysis of gene*group (0/1) interaction with outcome of 1Y visit APE score, covarying for age, baseline WRAT4 score, site, and baseline APE score. No genes attained genome-wide significance (p<5×10^−6^). B) Manhattan plot of TLC GWAS gene by group interaction associated with 1Y visit Learning and Memory (LM) cognitive domain score. GWAS genome-wide analysis of gene*group (0/1) interaction with outcome of 1Y LM score, covarying for age, baseline WRAT4 score, site, and baseline LM score. One gene, POC5 centriolar protein (*POC5*), attained genome-wide significance (p=1.99×10^−6^).

Gene analysis of LM domain performance identified one gene, POC5 centriolar protein (*POC5*), which was significantly enriched for variants associated with differences in cognitive performance in cases compared to controls (p=1.99×10^−6^, see Figure 5B for gene analysis results, supplementary Figure 2B for gene QQ plots).

Analyses performed with *APOE* e4 carrier status as an additional covariate did not differ significantly (data not shown).

## DISCUSSION

This first large-scale study of the association between genetic variation and longitudinal cognitive performance in older women with breast cancer and controls identified two novel loci and three genes of interest. The finding that women with cancer perform differently on cognitive assessments than controls based on minor allele carrier status suggests that genetic background may influence risk for CRCD, or may play a role in cognitive dysfunction following diagnosis and treatment.

These analyses identified two loci differentially associated with APE performance over time in cases and controls. The chromosome 1 SNP rs76859653 is intronic (non-coding) in the *HMCN1* gene. This gene encodes an extracellular protein of the immunoglobulin superfamily, suggesting that immune function may play a role in CRCD. The role in humans is unknown, but HMCN in *C. elegans* is involved in maintenance of cell polarity as well as cell migration and invasion^23^. Mutations in this gene have also been identified in gastric and colorectal cancers^24^. Interestingly, a mutation in *HMCN1* has been linked to occurrence of age-related macular degeneration^25,26^. Age-related macular degeneration occurs with an increased risk of dementia^27^, providing a potential connection between this gene and CRCD. Computational analysis of potential functions for this SNP including expression quantitative trait analyses did not reveal an obvious mechanism of SNP function; more work will be required to validate this finding and investigate the underlying molecular mechanisms for this locus in CRCD.

The second SNP identified in this analysis, rs78786199, occurs in an intergenic region of chromosome 2. The closest gene to this locus is forkhead box N2 (FOXN2), which is ubiquitously expressed and has been shown to suppress cancer proliferation and invasion^28^. Downregulation of this gene has been shown in acute myeloid leukemia, and was correlated with complex cytogenetic abnormalities^29^. Knockdown of this transcription factor was also associated with increased cancer cell proliferation, an impaired DNA damage response, and chromosomal instability^29^. While FOXN2 has not been specifically identified in neurodegenerative disease or cognitive functional research, perturbations in these molecular pathways have been identified in Alzheimer’s disease as well as cancer^30^. Additionally, rs78786199 is in a predicted binding site for several transcription factors, including Zinc Finger Imprinted 3, Interferon Regulatory Factors 1, 4, 7, and 8, Signal Transducer and Activator of Transcription 2, and Zinc Finger Protein 317. These transcription factors regulate numerous cellular processes, including hematopoiesis, inflammation, immune responses, cell proliferation, regulation of the cell cycle, and induction of growth arrest and programmed cell death in response to DNA damage, suggesting multiple mechanisms that could connect this locus with cancer and CRCD^31-36^. The location and current lack of validation for transcription factor binding at this intergenic locus makes functional interpretation challenging, however; more work is required to investigate and validate the molecular mechanisms underlying this locus.

Both SNPs show declining cognitive performance in cancer cases carrying minor alleles, in contrast to stable to improved performance in controls, suggesting that these loci increase risk for CRCD in older women who have experienced breast cancer and its treatment(s). Given that these loci were identified in a fairly homogeneous cohort of older White, female, well-educated breast cancer survivors, it seems reasonable to postulate that further study with larger, more diverse cohorts may uncover additional genetic etiologies for CRCD, providing tools for assessment of risk of cognitive decline in cancer survivors and/or avenues for therapeutic research.

Gene-level analysis identified *POC5*; this gene was enriched for variants differentially associated with LM performance change in cases and controls. There is evidence that the POC5 protein is involved in breast cancer cell proliferation and tumorigenesis^37^. POC5 is required for proper assembly of centrioles prior to cell division^38^. A study of histone deacetylases, dysregulation of which can result in carcinogenesis, showed that one mechanism of histone deacetylase action in cancer is to protect POC5 from degradation, resulting in cell cycle progression of cancer cells^37^. Mutations in *POC5* have been identified in adolescent idiopathic scoliosis (AIS); *in vitro* studies of these mutations have shown that they alter centrosome protein interactions, induce ciliary retraction, and impair cell-cycle progression^39^. A mutation in this gene has also been associated with retinitis pigmentosa^40^, an inherited form of retinal degeneration. Inherited retinal disease has extremely heterogeneous genetic etiologies, and syndromic forms can manifest with other symptoms including neurodegeneration, again providing a connection for this gene’s association to cognitive dysfunction as well as cancer^41^. However, much work remains to validate this finding and investigate the molecular mechanisms underlying *POC5* gene variants’ association with CRCD.

The TLC study is unique in having a larger number of individuals with genetic data, though still small in terms of genetics studies. Most studies of CRCD are too small to perform any genome-wide analyses, and are not powered to use for replication of rarer variants. Therefore, the ability to replicate this study is limited by the lack of well-powered studies of CRCD, particularly in older individuals who may be at increased risk. While there are a number of studies of older individuals with dementia that have cognitive data, these studies typically do not include well-documented cancer history or do not have sufficient populations of breast cancer survivors. These studies typically occur at much later time points following cancer diagnosis than a study of CRCD, making it difficult to meaningfully test for replication. An additional limitation was the exclusion of non-White participants to reduce population-driven genetic bias. This study was not powered to investigate the potential impact of genetic risk factors to CRCD in minority participants. It will be important for future studies to perform similar investigations in other racial and ethnic populations. Specifically, it is critical for additional studies to enroll larger cohorts of non-White participants, to enable the study of race-specific genetic factors underlying CRCD. Once enough samples/studies are available, it will be possible to perform meta-analyses including these data sets to investigate whether these or other genetic risk factors influence the risk for CRCD in minority populations.

These findings provide preliminary evidence that novel genetic loci may drive susceptibility to CRCD, and highlight a critical need for well-powered studies of the genetics of CRCD, particularly in older individuals who also have a greater risk for neurodegenerative disease and dementia. As more studies are funded to investigate this critical gap in understanding, it will be possible to perform meta-analyses with existing studies, similar to efforts to increase genetic sample size in Alzheimer’s and Parkinson’s disease research^42,43^. As larger studies become available, we expect meta-analyses to uncover additional genetic factors underlying CRCD, which may be used to further inform patient management and therapeutic research.

## Supporting information

STROBE checklist

## Data Availability

All data is available on request from the corresponding author.

## FIGURE LEGENDS

**Supplementary Figure 1. GWAS QQ Plots**. A) Attention, Processing speed, and Executive function (APE) QQ plot of –log10 p-values for GWAS analysis (APE domain one-year follow-up (1Y) visit ∼ SNP*Cancer case/control). B) Learning and Memory (LM) QQ plot of –log10 p-values for GWAS analysis (LM domain 1-year follow-up (1Y) visit ∼ SNP*cancer case/control). Dotted red lines depict 1:1 correlation of observed:expected p-values.

**Supplementary Figure 2. Gene QQ Plots**. A) Attention, Processing speed, and Executive function (APE) QQ plot of –log10 p-values for gene-level analysis (APE domain one-year follow-up (1Y) visit ∼ Gene*Cancer case/control). B) Learning and Memory (LM) QQ plot of – log10 p-values for gene analysis (LM domain 1Y follow-up (1Y) visit ∼ Gene*cancer case/control). Dotted red lines depict 1:1 correlation of observed:expected p-values.

## AUTHOR CONTRIBUTIONS

Conception and design: Jeanne S. Mandelblatt, Andrew J. Saykin, Tim A. Ahles, James C. Root

Provision of study materials or patients: Jeanne S. Mandelblatt, Tim A. Ahles, Andrew J. Saykin, Brenna C. McDonald, Deena M. A. Graham, Heather S. L. Jim, Paul B. Jacobsen, Sunita K. Patel

Collection and assembly of data: Wanting Zhai, Kelly N. H. Nudelman, Kwangsik Nho, Brent J. Small

Data analysis and interpretation: Kelly N. H. Nudelman, Kwangsik Nho, Michael Zhang, Brenna C. McDonald, Andrew J. Saykin, Jeanne S. Mandelblatt

Manuscript writing: Kelly N. H. Nudelman wrote first draft; all authors reviewed and edited

Final approval of manuscript: all authors

Accountable for all aspects of the work: all authors

## DATA AVAILABILITY STATEMENT

All data is available on request from the corresponding author.

## ACKNOWLEDGMENT

This research and the authors’ effort were supported by National Cancer Institute at the National Institutes of Health (NIH) grants R01CA129769 and R35CA197289 to JM. This study was also supported in part by National Institute on Aging at the National Institutes of Health grant R01AG068193 to JSM and AJS and by National Cancer Institute at the National Institutes of Health grant P30CA51008 to Georgetown Lombardi Comprehensive Cancer Center for support of the Biostatistics and Bioinformatics Resource and the Non-Therapeutic Shared Resource. KN was supported in part by National Institute on Aging, National Library of Medicine, and National Cancer Institute at the National Institutes of Health grants P30AG10133/P30AG072976. The work of AJS and BCM was supported in part by National Institute on Aging, National Library of Medicine, and National Cancer Institute at the National Institutes of Health grants P30AG10133/P30AG072976, R01AG19771, R01LM01136, R01CA244673, U01AG068057, and T32AG071444. TAA and JCR were supported in part by National Cancer Institute at the National Institutes of Health grants R01CA218496, R01CA172119, R03CA249548, and P30CA008748. TAA was also supported by U54CA137788. The work of JC was supported in part by the American Cancer Society Research Scholars grant 128660-RSG-15-187-01-PCSM and National Institute on Aging and National Cancer Institute at the National Institutes of Health grants R01CA237535 and R56AG068086. HJC was supported in part by the National Institute on Aging at the National Institutes of Health grant P30AG028716 for the Duke Pepper Center. SKP was supported in part by the American Cancer Society Research Scholars grant RSG-17-023-01-CPPB, and the National Institutes of Health grant R01CA261793.

## REFERENCES

1. Bluethmann SM, Mariotto AB, Rowland JH: Anticipating the “Silver Tsunami”: Prevalence Trajectories and Comorbidity Burden among Older Cancer Survivors in the United States. Cancer Epidemiol Biomarkers Prev 25:1029–36, 2016

2. Caswell-Jin JL, Plevritis SK, Tian L, et al: Change in Survival in Metastatic Breast Cancer with Treatment Advances: Meta-Analysis and Systematic Review. JNCI Cancer Spectr 2:pky062, 2018

3. Mandelblatt JS, Small BJ, Luta G, et al: Cancer-Related Cognitive Outcomes Among Older Breast Cancer Survivors in the Thinking and Living With Cancer Study. J Clin Oncol:JCO1800140, 2018

4. Mandelblatt JS, Jacobsen PB, Ahles T: Cognitive effects of cancer systemic therapy: implications for the care of older patients and survivors. J Clin Oncol 32:2617–26, 2014

5. Ahles TA, Li Y, McDonald BC, et al: Longitudinal assessment of cognitive changes associated with adjuvant treatment for breast cancer: the impact of APOE and smoking. Psychooncology 23:1382–90, 2014

6. Ahles TA, Saykin AJ, Noll WW, et al: The relationship of APOE genotype to neuropsychological performance in long-term cancer survivors treated with standard dose chemotherapy. Psychooncology 12:612–9, 2003

7. Carroll JE, Small BJ, Tometich DB, et al: Sleep disturbance and neurocognitive outcomes in older patients with breast cancer: Interaction with genotype. Cancer 125:4516–4524, 2019

8. Van Dyk K, Zhou X, Small BJ, et al: Protective Effects of APOE epsilon2 Genotype on Cognition in Older Breast Cancer Survivors: The Thinking and Living With Cancer Study. JNCI Cancer Spectr 5:pkab013, 2021

9. Wilkinson GS, Robertson GJ: Wide range achievement test (WRAT4). Lutz, FL, Psychological Assessment Resources, 2006

10. Folstein MF, Folstein SE, McHugh PR: “Mini-mental state”. A practical method for grading the cognitive state of patients for the clinician. J Psychiatr Res 12:189–98, 1975

11. Clapp JD, Luta G, Small BJ, et al: The Impact of Using Different Reference Populations on Measurement of Breast Cancer-Related Cognitive Impairment Rates. Arch Clin Neuropsychol 33:956–963, 2018

12. Van Dyk K, Zhou X, Small BJ, et al: Protective Effects of APOE epsilon2 Genotype on Cognition in Older Breast Cancer Survivors: The Thinking and Living With Cancer Study. JNCI Cancer Spectr 5, 2021

13. Purcell S, Neale B, Todd-Brown K, et al: PLINK: a tool set for whole-genome association and population-based linkage analyses. Am J Hum Genet 81:559–75, 2007

14. McCarthy S, Das S, Kretzschmar W, et al: A reference panel of 64,976 haplotypes for genotype imputation. Nat Genet 48:1279–83, 2016

15. Das S, Forer L, Schonherr S, et al: Next-generation genotype imputation service and methods. Nat Genet 48:1284–1287, 2016

16. Watanabe K, Taskesen E, van Bochoven A, et al: Functional mapping and annotation of genetic associations with FUMA. Nat Commun 8:1826, 2017

17. de Leeuw CA, Mooij JM, Heskes T, et al: MAGMA: generalized gene-set analysis of GWAS data. PLoS Comput Biol 11:e1004219, 2015

18. Watanabe K, Umicevic Mirkov M, de Leeuw CA, et al: Genetic mapping of cell type specificity for complex traits. Nat Commun 10:3222, 2019

19. Rentzsch P, Witten D, Cooper GM, et al: CADD: predicting the deleteriousness of variants throughout the human genome. Nucleic Acids Res 47:D886–D894, 2019

20. Boyle AP, Hong EL, Hariharan M, et al: Annotation of functional variation in personal genomes using RegulomeDB. Genome Res 22:1790–7, 2012

21. GTEx Consortium: The GTEx Consortium atlas of genetic regulatory effects across human tissues. Science 369:1318–1330, 2020

22. Castro-Mondragon JA, Riudavets-Puig R, Rauluseviciute I, et al: JASPAR 2022: the 9th release of the open-access database of transcription factor binding profiles. Nucleic Acids Res 50:D165–D173, 2022

23. Vogel BE, Muriel JM, Dong C, et al: Hemicentins: what have we learned from worms? Cell Res 16:872–8, 2006

24. Lee SH, Je EM, Yoo NJ, et al: HMCN1, a cell polarity-related gene, is somatically mutated in gastric and colorectal cancers. Pathol Oncol Res 21:847–8, 2015

25. Schultz DW, Klein ML, Humpert AJ, et al: Analysis of the ARMD1 locus: evidence that a mutation in HEMICENTIN-1 is associated with age-related macular degeneration in a large family. Hum Mol Genet 12:3315–23, 2003

26. McKay GJ, Clarke S, Hughes A, et al: A novel diagnostic test detects a low frequency of the hemicentin Gln5345Arg variant among Northern Irish age related macular degeneration patients. Mol Vis 10:682–7, 2004

27. Shang X, Zhu Z, Huang Y, et al: Associations of ophthalmic and systemic conditions with incident dementia in the UK Biobank. Br J Ophthalmol, 2021

28. Liu XH, Liu LP, Xu XM, et al: FOXN2 suppresses the proliferation and invasion of human hepatocellular carcinoma cells. Eur Rev Med Pharmacol Sci 25:731–737, 2021

29. Cheng CK, Chan NP, Wan TS, et al: Helicase-like transcription factor is a RUNX1 target whose downregulation promotes genomic instability and correlates with complex cytogenetic features in acute myeloid leukemia. Haematologica 101:448–57, 2016

30. Nudelman KNH, McDonald BC, Lahiri DK, et al: Biological Hallmarks of Cancer in Alzheimer’s Disease. Mol Neurobiol 56:7173–7187, 2019

31. Negishi H, Taniguchi T, Yanai H: The Interferon (IFN) Class of Cytokines and the IFN Regulatory Factor (IRF) Transcription Factor Family. Cold Spring Harb Perspect Biol 10, 2018

32. Xia X, Wang W, Yin K, et al: Interferon regulatory factor 8 governs myeloid cell development. Cytokine Growth Factor Rev 55:48–57, 2020

33. Chen YJ, Luo SN, Dong L, et al: Interferon regulatory factor family influences tumor immunity and prognosis of patients with colorectal cancer. J Transl Med 19:379, 2021

34. Dornan D, Eckert M, Wallace M, et al: Interferon regulatory factor 1 binding to p300 stimulates DNA-dependent acetylation of p53. Mol Cell Biol 24:10083–98, 2004

35. Bowie ML, Ibarra C, Seewalt VL: IRF-1 promotes apoptosis in p53-damaged basal-type human mammary epithelial cells: a model for early basal-type mammary carcinogenesis. Adv Exp Med Biol 617:367–74, 2008

36. Bluyssen HA, Levy DE: Stat2 is a transcriptional activator that requires sequence-specific contacts provided by stat1 and p48 for stable interaction with DNA. J Biol Chem 272:4600–5, 1997

37. Wei H, Ma W, Lu X, et al: KDELR2 promotes breast cancer proliferation via HDAC3-mediated cell cycle progression. Cancer Commun (Lond) 41:904–920, 2021

38. Azimzadeh J, Hergert P, Delouvee A, et al: hPOC5 is a centrin-binding protein required for assembly of full-length centrioles. J Cell Biol 185:101–14, 2009

39. Hassan A, Parent S, Mathieu H, et al: Adolescent idiopathic scoliosis associated POC5 mutation impairs cell cycle, cilia length and centrosome protein interactions. PLoS One 14:e0213269, 2019

40. Weisz Hubshman M, Broekman S, van Wijk E, et al: Whole-exome sequencing reveals POC5 as a novel gene associated with autosomal recessive retinitis pigmentosa. Hum Mol Genet 27:614–624, 2018

41. Tatour Y, Ben-Yosef T: Syndromic Inherited Retinal Diseases: Genetic, Clinical and Diagnostic Aspects. Diagnostics (Basel) 10, 2020

42. Kunkle BW, Grenier-Boley B, Sims R, et al: Genetic meta-analysis of diagnosed Alzheimer’s disease identifies new risk loci and implicates Abeta, tau, immunity and lipid processing. Nat Genet 51:414–430, 2019

43. Nalls MA, Pankratz N, Lill CM, et al: Large-scale meta-analysis of genome-wide association data identifies six new risk loci for Parkinson’s disease. Nat Genet 46:989–93, 2014

