## Supplementary Methods for "Genetic variants associated with longitudinal cognitive performance in older breast cancer patients and controls"

**Data Collection**

*Neuropsychological Testing*

The APE domain score includes the Digits Forward and Backward subtests from the Neuropsychological Assessment Battery (NAB), Trail Making Tests A and B, the Controlled Oral Word Association Test, and the Digit Symbol subtest from the Wechsler Adult Intelligence Scale-III^1-5^. The LM domain includes the Logical Memory I and II subtests from the Weschler Memory Scale-III and the Immediate Recall, Short Delayed Recall, and the Long Delayed Recall scores from the NAB List Learning Test^1,4,5^.

*Genetics*

GWAS quality control included genetic sex check (all female), removal of subjects with >10% SNP failure rate, removal of SNPs with >5% failure rate, removal of duplicate SNPs and those with minor allele frequencies <1%, and removal of SNPs with Hardy-Weinberg equilibrium (HWE) p<1x10^-6^. SNPs that validate the assumptions of Hardy-Weinberg equilibrium were removed, as if SNPs have significantly different numbers of homozygous and heterozygous carriers based on the SNP population allele frequency, it could indicate genotyping errors. Identity-by-descent, which evaluates the percentage of regions of identical DNA shared between two individuals to determine relatedness, was used to check for relationships among participants.

Each batch of genetic data was then merged with HapMap3 genotype data representing distinct genetic ancestral reference populations, and analyzed with multidimensional scaling (MDS) in PLINK^6^. Multidimensional scaling is used to evaluate the clustering of genetic data for TLC cases and controls with the reference population genetic data, to identify the genetic ancestry of each participant. Principal components were calculated for genetic ancestry using the clustering algorithm, and participants of white non-Hispanic ancestry were selected for imputation.

Imputed data was further filtered to remove SNPs with HWE p<1x10^-6^, as well as SNPs with minor allele frequencies <1%.

**Statistical Methods and Analyses**

The Functional Mapping and Annotation of Genome-wide Association Studies (FUMA GWAS) program^7^ v1.3.6a. was used to create Manhattan and Q-Q plots, and identify significant SNPs. Manhattan plots visualize GWAS results across the genome (x-axis), showing each SNP result as a separate point by location and –log_10_ p-value (y-axis). Q-Q plots visualize the observed vs. expected –log_10_ p-value for SNPs to detect whether there is any unaccounted-for population structural bias (significant deviation from a linear trend line, also indicated by genome inflation calculation). The genomic inflation factor for observed vs. expected test statistics was calculated separately in R v3.6^8^.

**References**

1.	Stern R, White T: Neuropsychological Assessment Battery: Administration, Scoring, and Interpretation. Lutz, FL, Psychological Assessment Resources, Inc., 2003

2.	Reitan R: Validity of the Trail Making Test as an indicator of organic brain damage. Perceptual and motor skills 8:271-276, 1958

3.	Patterson J: Controlled Oral Word Association Test, in Kreutzer J, DeLuca J, Caplan B (eds): Encylopedia of Clinical Neuropsychology. New York, NY, Springer New York, 2011, pp 703-706

4.	Wechsler D: WAIS-III, Wechsler Adult Intelligence Scale: Administration and Scoring Manual. San Antonio, TX, Psychological Corporation, 1997

5.	Wechsler D: Manual for the Wechsler Memory Scale-Revised. San Antonio, TX, Psychological Corporation, 1987

6.	Purcell S, Neale B, Todd-Brown K, et al: PLINK: a tool set for whole-genome association and population-based linkage analyses. Am J Hum Genet 81:559-75, 2007

7.	Watanabe K, Taskesen E, van Bochoven A, et al: Functional mapping and annotation of genetic associations with FUMA. Nat Commun 8:1826, 2017

8.	R Development Core Team: R: a language and environment for statistical computing. Vienna, Austria, R Foundation for Statistical Computing, 2019
