## Supplementary Figure 1 for "Genetic variants associated with longitudinal cognitive performance in older breast cancer patients and controls"

### Slide 1
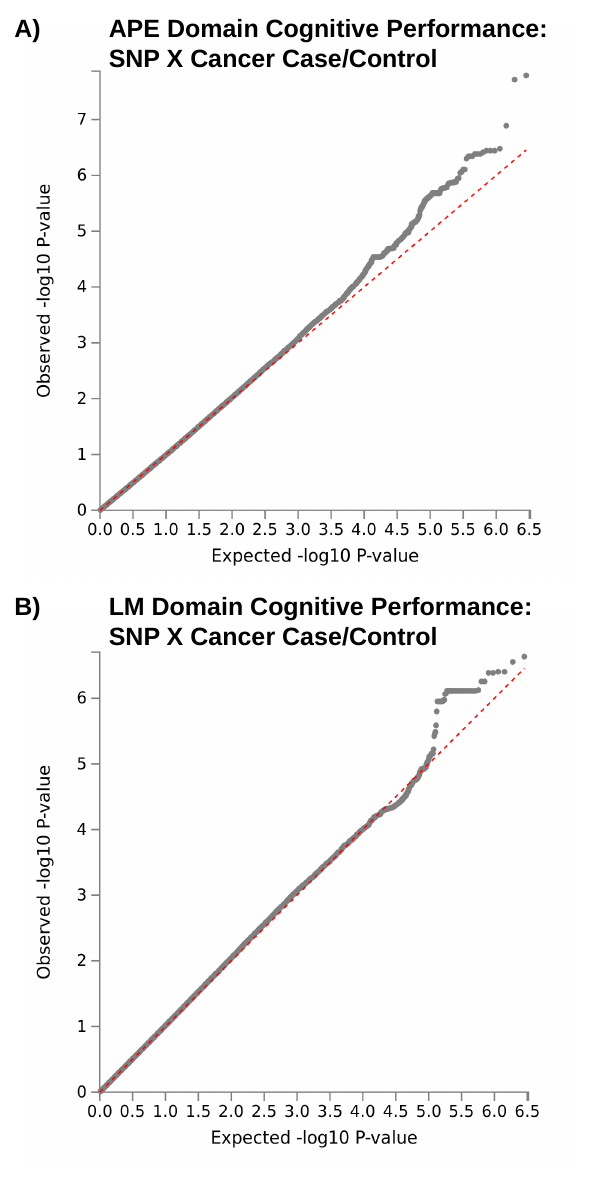

A)
APE Domain Cognitive Performance: SNP X Cancer Case/Control
B)
LM Domain Cognitive Performance: SNP X Cancer Case/Control
